# COVID-19 related mortality and spread of disease in long-term care: a living systematic review of emerging evidence

**DOI:** 10.1101/2020.06.09.20125237

**Authors:** Maximilian Salcher-Konrad, Arnoupe Jhass, Huseyin Naci, Marselia Tan, Yousef El-Tawil, Adelina Comas-Herrera

## Abstract

**Background:** Policy responses to mitigate the impact of the COVID-19 pandemic on long-term care (LTC) require robust and timely evidence on mortality and spread of the disease in these settings. The aim of this living systematic review is to synthesise early international evidence on mortality rates and incidence of COVID-19 among people who use and provide LTC.

**Methods:** We report findings of a living systematic review (CRD42020183557), including studies identified through database searches up to 26 June 2020. We searched seven databases (MEDLINE; Embase; CINAHL Plus; Web of Science; Global Health; WHO COVID-19 Research Database; medRxiv) to identify all studies reporting primary data on COVID-19 related mortality and incidence of disease among LTC users and staff. We excluded studies not focusing on LTC. Included studies were critically appraised and results on number of deaths and COVID-19 related mortality rates, case fatality rates, and excess deaths (co-primary outcomes), as well as incidence of disease, hospitalisations, and ICU admissions were synthesised narratively.

**Findings:** A total of 54 study reports for 49 unique primary studies or outbreak reports were included. Outbreak investigations in LTC facilities found COVID-19 incidence rates of between 0.0% and 71.7% among residents and between 0.4% and 64.0% among staff at affected facilities. Mortality rates varied from 0.0% to 17.1% of all residents at outbreak facilities, with case fatality rates between 0.0% and 33.7%. In included studies of outbreaks, no LTC staff members had died.

Studies of wider LTC populations found that between 0.4% and 40.8% of users, and between 4.0% and 23.8% of staff were infected, although the generalisability of these studies is limited.

There was limited information on the impact of COVID-19 on LTC in the community.

**Interpretation:** Long-term care users have been particularly vulnerable to the COVID-19 pandemic. However, we found wide variation in spread of disease and mortality rates between outbreaks at individual LTC facilities. Further research into the factors determining successful prevention and containment of COVID-19 outbreaks is needed to protect long-term care users and staff.

**Funding:** This work was partially conducted as part of the “Strengthening responses to dementia in developing countries” (STRiDE) project, supported by the UK Research and Innovation’s Global Challenges Research Fund (ES/P010938/1). The funders had no role in the design and execution of this study, interpretation of its results, and decision to submit this work to be published.

## Introduction

The coronavirus disease 2019 (COVID-19) pandemic has taken a substantial morbidity and mortality toll on the world.^1^ Over the course of the evolving pandemic, public attention in some countries has shifted towards long-term care facilities as “ground zero”.^2^ Early evidence on risk factors for severe outcomes suggested that residents of long-term care facilities, such as nursing homes and residential care facilities for people who need medical support or support in their activities of daily living, may be particularly vulnerable. Studies have shown that older people and those with underlying health conditions, including hypertension, diabetes, cardiovascular disease, chronic lung disease, obesity, and cancer, are more likely to experience severe outcomes after contracting the disease.^3^ Importantly, case series from China, Italy, the United States (US), and the United Kingdom (UK), have shown higher mortality rates among older people who contract COVID-19.^4,5,6,7^ Official figures showed that a substantial proportion of COVID-19 related deaths – more than 50% of all deaths in many high-income countries – is concentrated among long-term care users.^8^ While older people and those with chronic conditions would already have higher mortality rates in the absence of a pandemic, a modelling study for the United Kingdom has shown that excess deaths due to COVID-19 are likely to be concentrated among older people.^9^ It has also been suggested that older age and some chronic conditions were associated with an increased risk of infection with SARS-CoV-2, the virus causing COVID-19.^8^

While the combination of a population of older people with underlying health conditions living in close proximity to each other suggests the long-term care sector to be at particularly high risk, specific evidence on COVID-19 infections and associated deaths in this setting was initially slow to emerge. An early rapid review on deaths in care homes conducted in mid-April 2020 identified only three studies on infection rates and COVID-19 incidence and mortality in long-term care homes (all from the US).^8^ These studies showed wide variation in the proportion of residents and staff being infected (with the majority of people who contracted COVID-19 asymptomatic at the time of testing), in the spread of the disease between different care homes, and in case fatality rates among nursing home residents, which were reported to be as high as 33%.^10,11,12^ As the pandemic continues to spread, more evidence about the spread and impact of COVID-19 in long-term care settings is emerging, including outbreak reports and studies about infection rates and outcomes among those receiving long-term care services (long-term care users) and those providing them (long-term care staff), including both for institutional settings and community-based services. Indeed, the number of records in PubMed retrieved through a combination of search terms for COVID-19 and long-term care increased by approximately 100 records per week from the end of April to the end of June.

Given the vulnerability of the population relying on long-term care services and the potentially large burden of COVID-19 in this sector, timely and evidence-based policy responses are required. We therefore aimed to systematically collate and synthesise available and newly emerging evidence on the number of long-term care users and staff who contract COVID-19 and experience severe outcomes, including death, and the spread of disease in long-term care settings.

## Methods

We conducted a systematic review of available evidence on COVID-19 infection rates and mortality among users and providers of long-term care services (PROSPERO: CRD42020183557). Due to the rapidly evolving nature of the situation and an expected increase in research focusing on COVID-19 in long-term care, database searches will be updated continuously, and findings incorporated as a living systematic review. The reporting of this review is PRISMA-compliant (see supplemental file).^13^

### Search Strategy & Selection Criteria

Potentially eligible studies were identified through systematic searches of seven electronic databases (MEDLINE, Embase, CINAHL plus, Web of Science, Global Health, the World Health Organization’s COVID-19 Research Database, medRxiv). Search terms were based on published search blocks for COVID-19 related studies and were adapted to each database (see supplemental file).^14,15^ We included full study reports and research letters published in peer-reviewed journals or on pre-print servers since 1 January 2020 in order to capture newly emerging evidence pertinent to the COVID-19 pandemic. Initial database searches were conducted on 15 May 2020 and updated weekly up to 26 June 2020.

Inclusion criteria were defined following the CoCoPop (Condition, Context and Population) framework, as recommended by the Joanna Briggs Institute for systematic reviews of prevalence and incidence.^16^ Studies were eligible for inclusion if they reported primary data on COVID-19 related mortality (including mortality rate among the population of interest, case fatality rate (CFR), and excess deaths compared to previous periods) or spread of COVID-19 among users and staff of long-term care services. Long-term care services included both institutional and community (i.e., care provided in the homes of patients) settings. We excluded studies that focused on COVID-19 mortality and infection rates in non-long-term care settings, studies of infectious disease outbreaks other than COVID-19, modelling studies, as well as opinion pieces and review articles that did not report original data. We had previously included reports of official figures of COVID-19 deaths and the proportion of long-term care users among them but decided to exclude these figures as more evidence from primary research studies became available. Official figures are summarised by members of our group in separate reports.^8^

Title and abstract screening, as well as full text review was undertaken by three reviewers (AJ, MS-K, and MT). To ensure consistency, all studies deemed eligible for inclusion were again reviewed by one reviewer (MS-K). Records reporting on the same study or outbreak were combined.

### Data Extraction & Synthesis

A standardised template was used to extract data at the study level, including information on study design; care setting (institutional vs. community); how COVID-19 was diagnosed and confirmed; baseline characteristics of participants; absolute number of deaths and mortality rates for people with confirmed and suspected COVID-19; CFRs; excess deaths; absolute numbers and rates of confirmed and suspected COVID-19; and rates of hospitalisation and intensive care unit (ICU) admissions among people with confirmed and suspected COVID-19. All study participant characteristics and outcomes data were extracted separately for long-term care users and staff. For rates, we also recorded how numerator and denominator were defined, as well as the follow-up time over which outcomes were measured.

Based on extracted data, we calculated the mortality rate directly attributable to COVID-19 (all deaths among those who contracted COVID-19/all long-term care users or staff), CFR (all deaths among those who contracted COVID-19/all long-term care users or staff who contracted COVID-19), incidence of COVID-19 (all those who contracted COVID-19/all long-term care users or staff), and incidence of hospital and ICU admissions (all hospital or ICU admissions/all long-term care users or staff who contracted COVID-19). Due to heterogeneity in the definitions of numerators, denominators, and follow-up times across included studies, data were not pooled. Instead, results are summarised narratively and presented in tables, including information on sample characteristics, follow-up time, and case definitions, as appropriate. Where studies reported on overlapping populations, we gave preference to those with larger sample sizes and longer follow-up times.

In addition to the pre-specified outcomes above, we extracted information on the proportion of asymptomatic people with COVID-19 at time of testing, and findings of studies comparing outcomes in long-term care users to others.

### Critical Appraisal

The quality of included studies reporting figures relating to mortality rates, CFR, or disease incidence were assessed using the Joanna Briggs Institute critical appraisal tool for prevalence studies.^17^ The tool includes nine questions about the appropriateness of the sampling frame, sampling of participants, sample size, description of study setting and participants, data analysis, identification of cases, measure of disease, statistical analysis, and response rate. We summarised appraisals as the number of items that were deemed appropriate for each study.

We did not assess risk of bias across studies.

## Results

The first report of this living systematic review was published on 9 June 2020,^18^ and an updated version on 29 June 2020. Since then, 21 additional studies were included, leading to a total of 54 study reports for 49 unique studies or outbreak reports (Figure 1).^7,10,11,12,19,20,21,22,23, 24,25,26,27,28,29,30,31,32,33,34,35,36,37,38,39,40,41,42,43,44,45,46,47,48,49,50,51,52,53,54,55,56,57,58,59,60,61,62,63,64,65,66,67,68^

**Figure 1:**
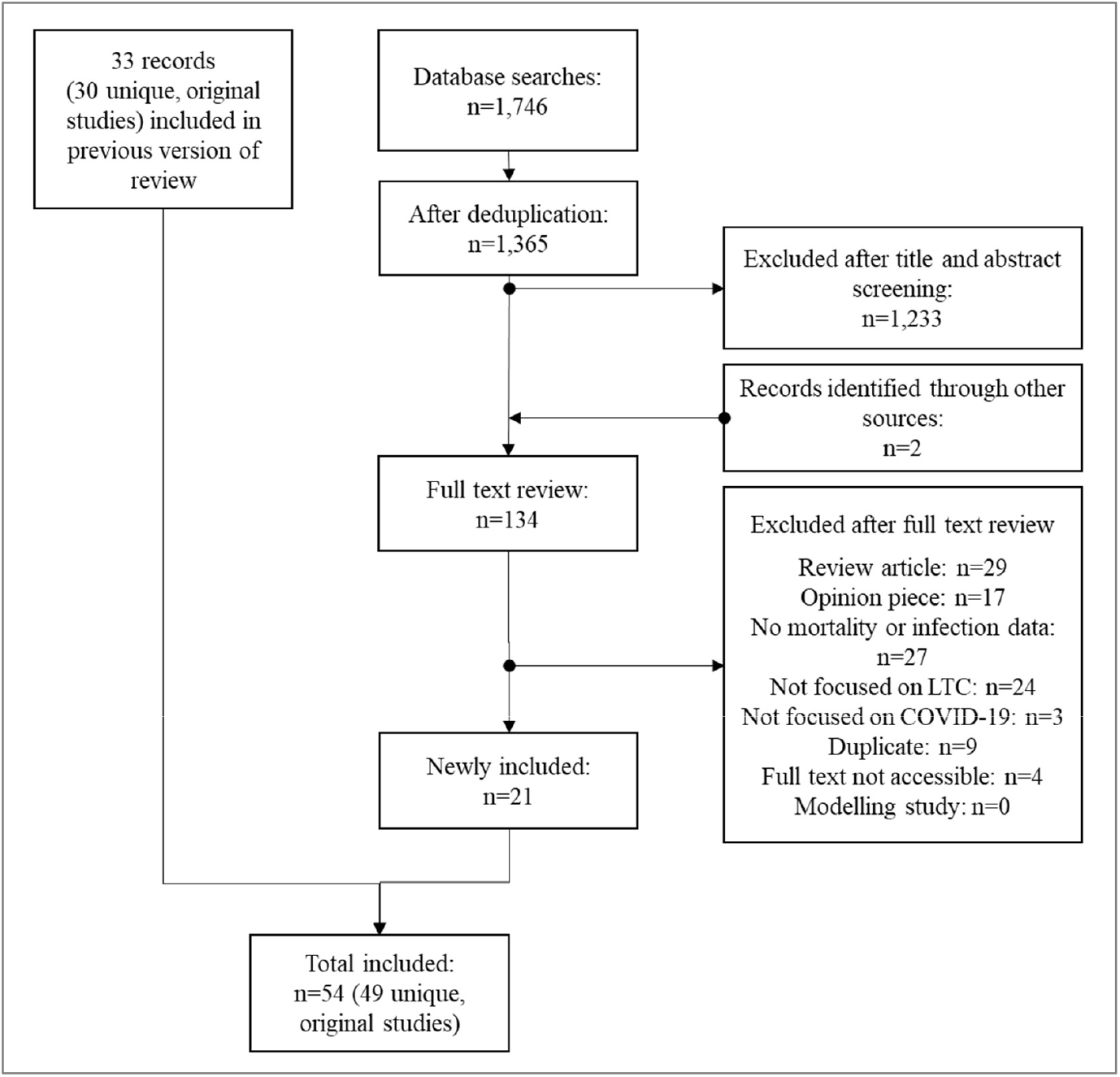
Flow chart for selection of included studies.

An overview of study characteristics is provided in Table 1. Twenty reported on individual outbreaks, while the rest reported on wider populations. Included studies were conducted in 14 countries (20 in the United States, six each in Spain and the United Kingdom, three each in Canada and South Korea, two each in Belgium and France, and one each in Germany, Hong Kong, Hungary, Ireland, Israel, the Netherlands, and Poland). All included studies except for three were exclusively conducted in institutional care settings. Three studies reported on home-based or community-based care.

**Table 1:**
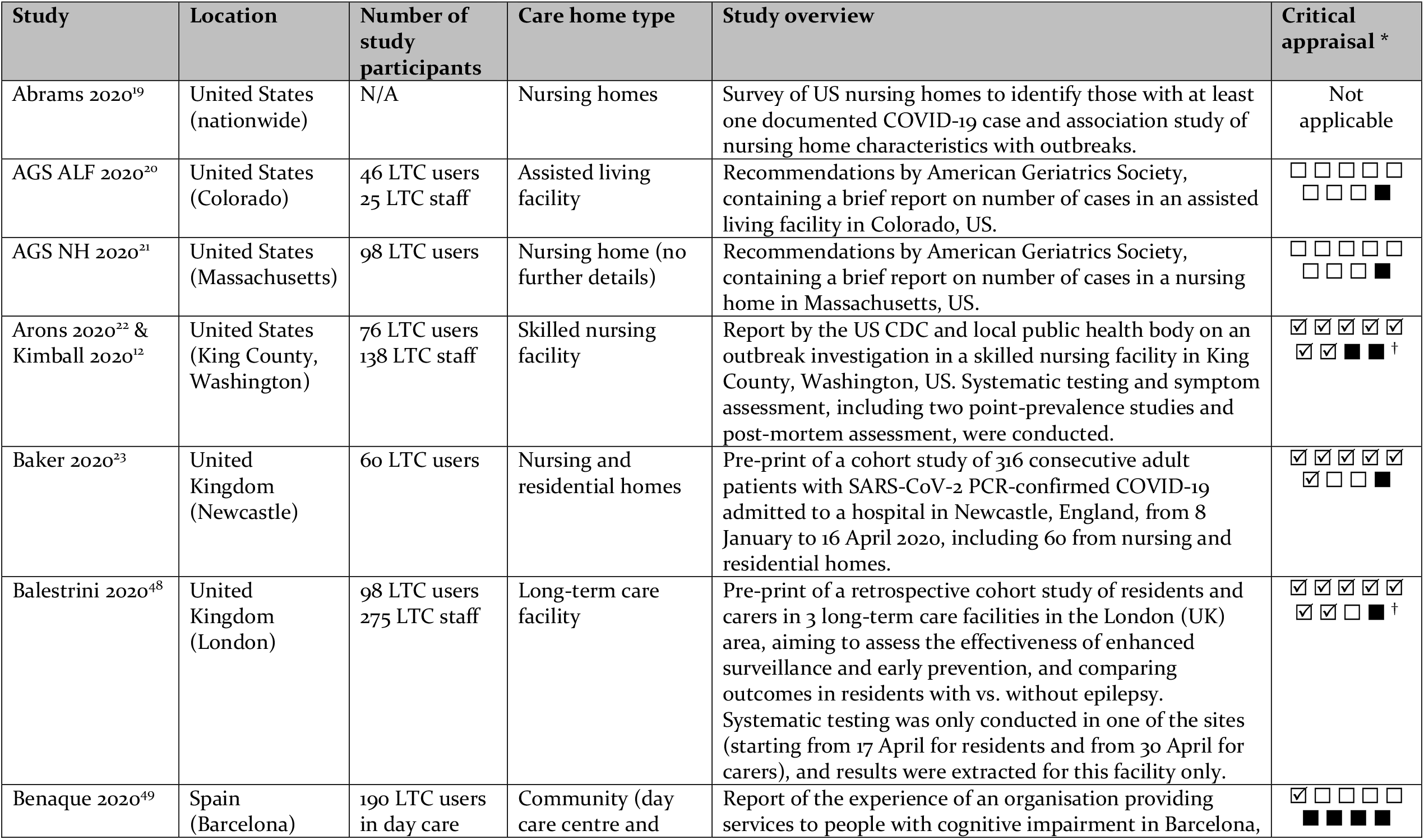
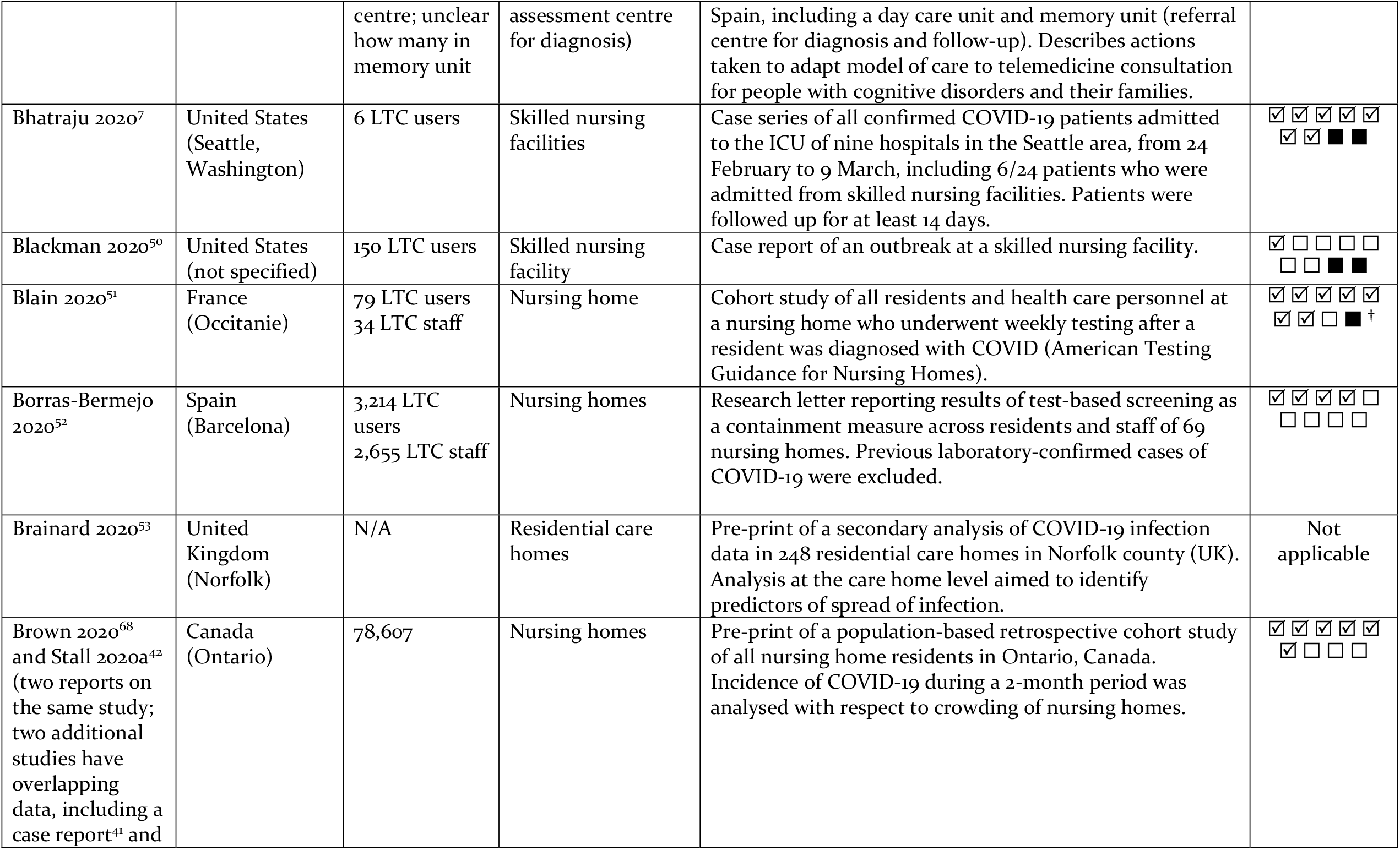
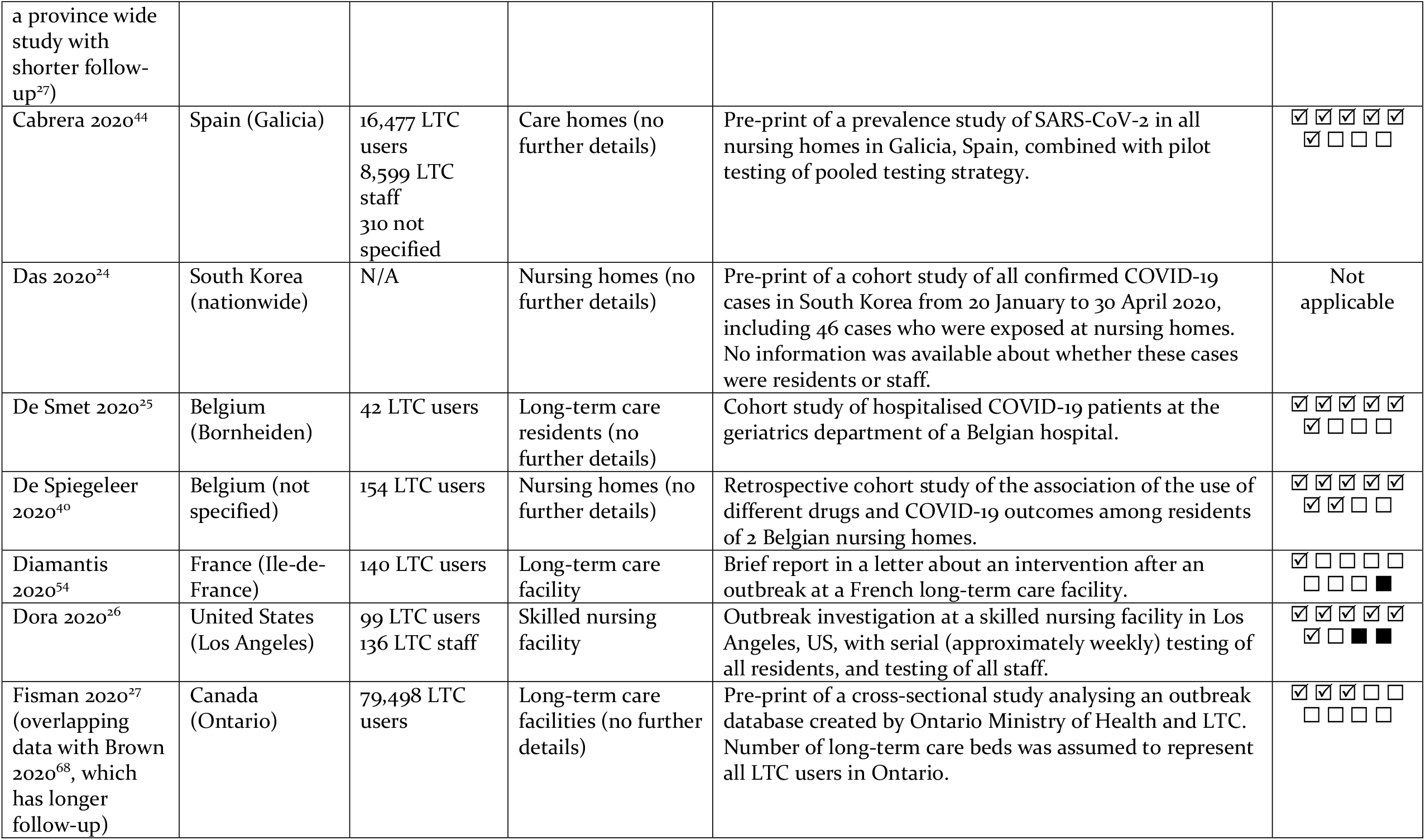
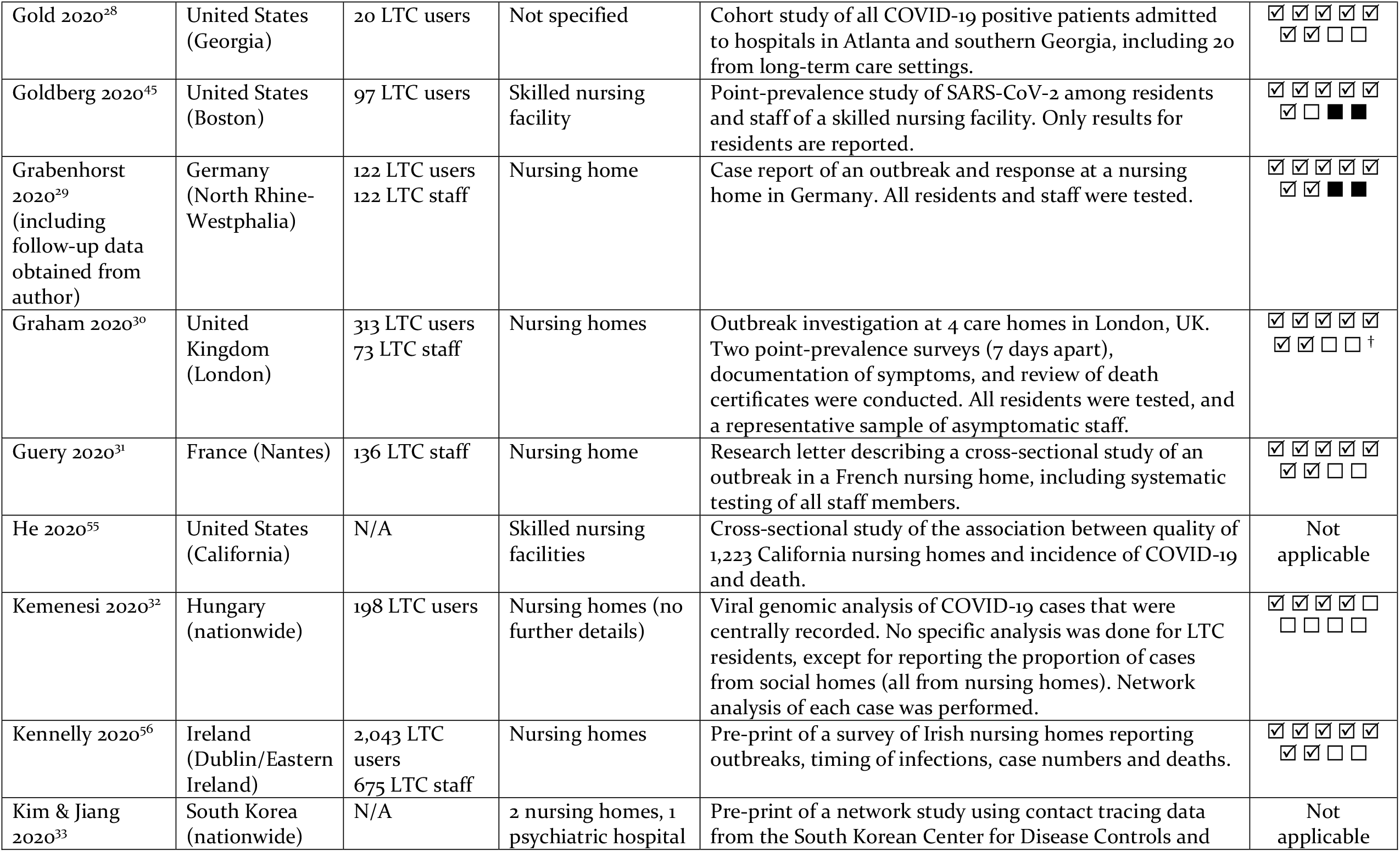
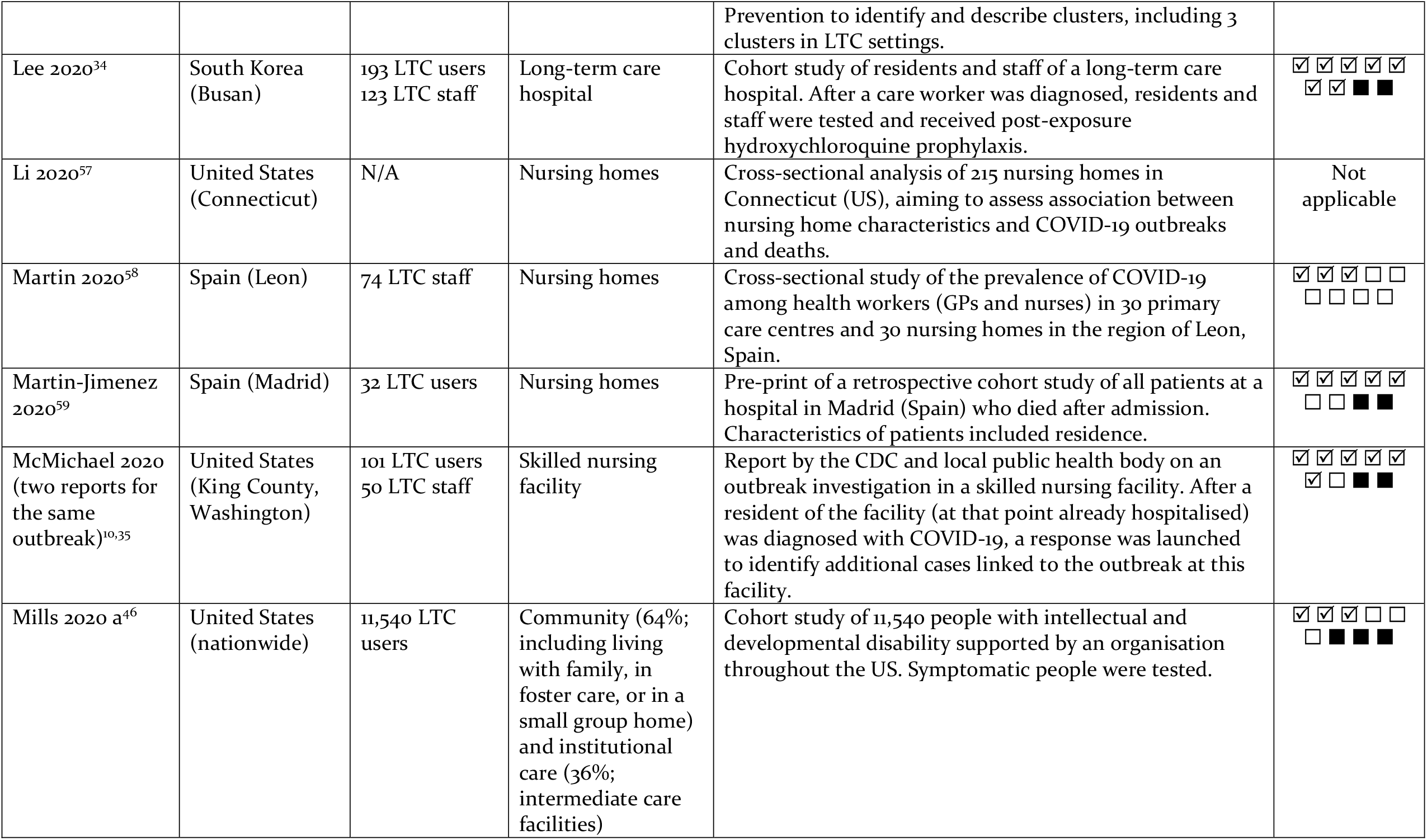
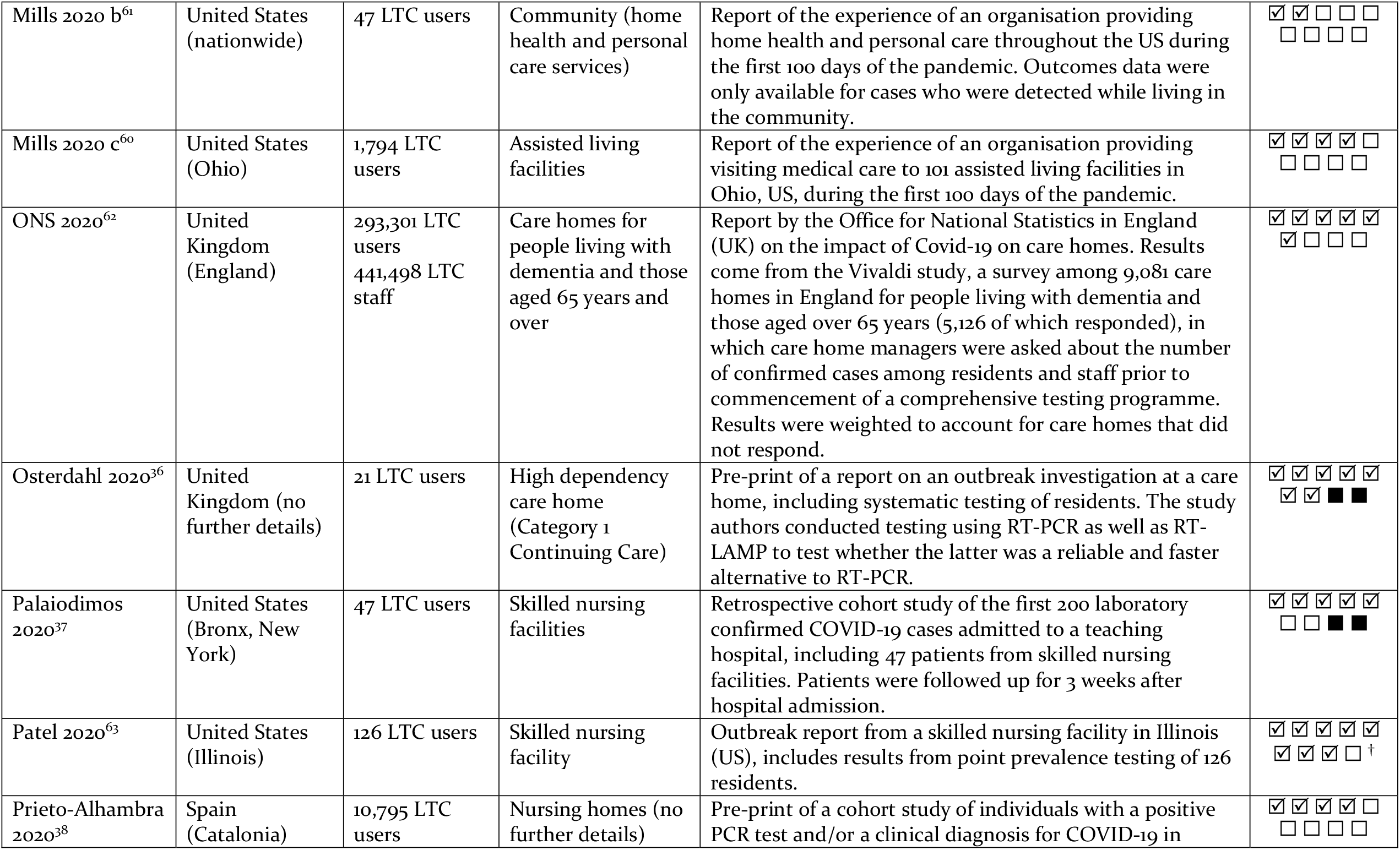
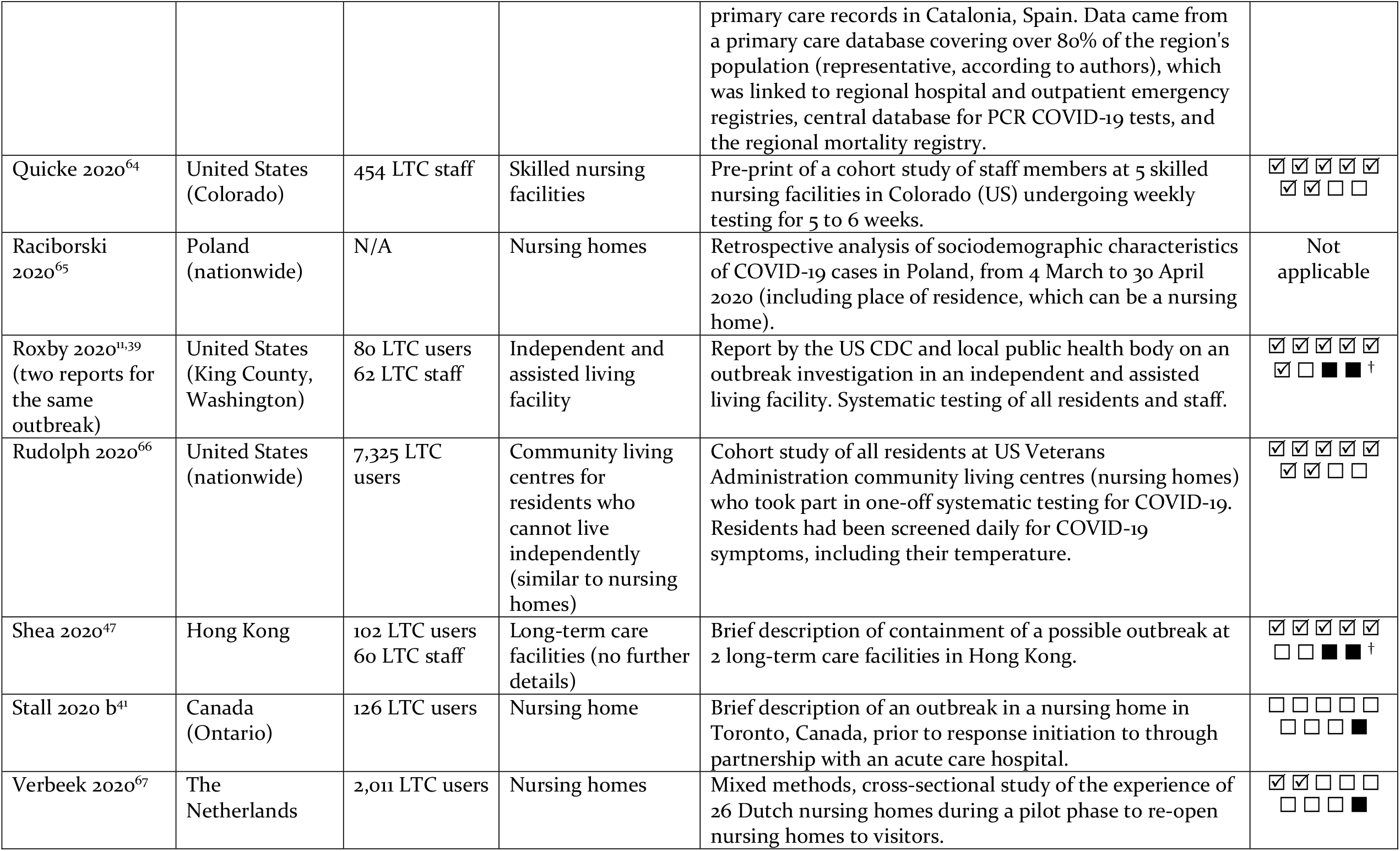
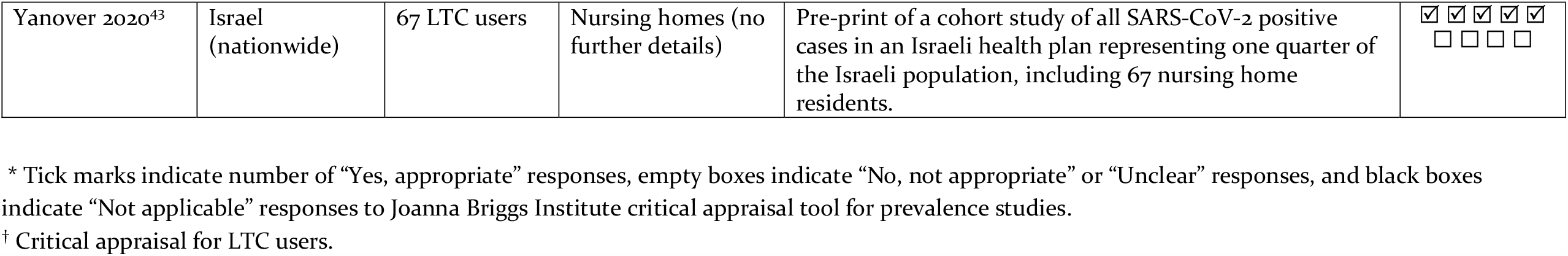
Overview of included studies.

### Evidence on disease incidence in institutionalised long-term care settings

Evidence on the spread of disease within institutional long-term care settings was available from 25 studies,^20,21,22,26,29,30,31,34,36,39,41,45,47,48,51,52,56,58,60,62,63,64,66,67,68^ including 17 studies reporting the number of people who contracted COVID-19 in facilities facing potential outbreaks (Table 2) and eight studies reporting relevant figures among regional or national populations of long-term care facilities (Table 3).

**Table 2:** Incidence of confirmed COVID-19 among long-term care users and staff in potential outbreak facilities.

| Study | Incidence rate | Number of people who contracted COVID-19 | Number of users / staff | Time period | Source population | Diagnosis |
| --- | --- | --- | --- | --- | --- | --- |
| **Incidence of confirmed COVID-19 among long-term care users in potential outbreak facilities** |  |  |  |  |  |  |
| AGS ALF 2020 (US) | 71.7% | 33 | 46 | Not reported | No details reported | Positive test (no details) |
| Stall 2020 b (Canada) | 70.6% | 89 | 126 | 2 weeks | All nursing home residents, assuming full occupancy of 126-bed facility | Infected (no details) |
| Arons 2020 (US) | 63.2% | 48 | 76 | 7 days | 76 out of all 89 residents at the investigated skilled nursing facility | PCR |
| Goldberg 2020 (US) | 53.6% | 52 | 97 | Not reported (point-prevalence survey) | All 97 nursing home residents were tested | PCR |
| AGS NH 2020 (US) | 51.0% | 50 | 98 | Not reported | All residents at the investigated nursing home who were asymptomatic | Positive test (no details) |
| Blain 2020 (France) | 48.1% | 38 | 79 | 6 weeks | All 79 residents at the nursing home participating in weekly PCR testing | PCR |
| Osterdahl 2020 (UK) | 47.6% | 10 | 21 | 4 days | 21 out of all 24 residents at the investigated nursing home | PCR |
| Graham 2020 (UK) | 40.3% | 126 | 313 | 7 days | Appr. 94% of all residents at the time of systematic testing (available and consented to testing) | PCR |
| Patel 2020 (US) | 26.2% | 33 | 126 | Point prevalence | 126 residents at the investigated facility who consented to testing (1 refused) | PCR |
| Dora 2020 (US) | 19.2% | 19 | 99 | 26 days | All 99 residents at the facility at the time of outbreak | PCR |
| Grabenhorst 2020 (Germany) | 13.1% | 16 | 122 | 69 days | All 122 residents at the time of systematic testing | PCR |
| Balestrini 2020 (UK) | 9.2% | 9 | 98 | 12 weeks | All 98 residents at the long-term care facility | PCR |
| Roxby 2020 (US) | 5.0% | 4 | 80 | 7 days | All residents at the investigated assisted living facility except for 2 index cases | PCR |
| Shea 2020 (Hong Kong) | 0.0% | 0 | 102 | 28 days | Most residents at 2 long-term care facilities where an infected nurse worked (total number of residents at the facilities is unclear) | PCR |
| Lee 2020 (Korea) | 0.0% | 0 | 193 | 20 days | All 193 inpatients at the investigated long-term care hospital who were exposed to an infected care worker | PCR |
| **Incidence of confirmed COVID-19 among long-term care staff in potential outbreak facilities** |  |  |  |  |  |  |
| AGS ALF 2020 (US) | 64.0% | 16 | 25 | Not reported | No details reported | Positive test (no details) |
| Patel 2020 (US) | 45.2% | 19 | 42 | Point prevalence | 42 staff members were tested out of 70 who were offered testing | PCR |
| Blain 2020 (France) | 23.5% | 8 | 34 | 6 weeks | All 35 health care personnel at the nursing home participating in weekly PCR testing | PCR |
| Quicke 2020 (US) | 19.9% | 70 | 351 | 6 weeks | 351 workers at four facilities where cases were detected (systematic, weekly testing among consenting individuals) | PCR |
| Arons 2020 (US) | 18.8% | 26 | 138 | 24 days | All 138 full-time staff members at the investigated skilled nursing facility (51 of which were tested) | PCR |
| Dora 2020 (US) | 5.9% | 26 | 138 | 13 days | All 138 full-time staff members at the investigated skilled nursing facility (51 of which were tested) | PCR |
| Graham 2020 (UK) | 4.1% | 3 | 73 | 1-2 days | Sample of 11.8% of staff members asymptomatic at the time of testing (representing all staff roles including health care assistants, registered nurses, kitchen staff, administrators, domestic and maintenance staff) | PCR |
| Grabenhorst 2020 (Germany) | 3.3% | 4 | 122 | 69 days | All 122 staff members at the time of systematic testing (including maintenance personnel) | PCR |
| Roxby 2020 (US) | 3.2% | 2 | 62 | 2 days | All staff members working at the investigated facility | PCR |
| Guery 2020 (France) | 2.2% | 3 | 136 | 2 days | All 136 staff members, health workers, and administrative personnel at the investigated nursing home | PCR |
| Shea 2020 (Hong Kong) | 1.7% | 1 | 60 | 28 days | All 60 staff at 2 long-term care facilities where an infected nurse worked | PCR |
| Lee 2020 (Korea) | 1.5% | 2 | 132 | 20 days | All 123 staff at the investigated long-term care hospital | PCR |
| Balestrini 2020 (UK) | 0.4% | 1 | 275 | 12 weeks | All 275 asymptomatic carers at the facility (systematic testing was started 4 weeks after the first case was detected) | PCR |

Abbreviations: PCR, polymerase chain reaction

**Table 3:** Incidence of confirmed COVID-19 among long-term care users and staff (population-wide studies)

| Study | Incidence rate | Number of people who contracted COVID-19 | Number of users / staff | Time period | Source population | Diagnosis |
| --- | --- | --- | --- | --- | --- | --- |
| **Incidence of confirmed COVID-19 among long-term care users (population-wide studies)** |  |  |  |  |  |  |
| Kennelly 2020 (Dublin and Eastern Ireland, Ireland) | 40.8% | 710 | 1741 | 12 weeks | All 1741 residents in nursing homes which had outbreaks and responded to survey (excluding nursing homes without outbreaks). | PCR |
| Borras-Bermejo 2020 (Barcelona, Spain) | 23.9% | N/A | 3214 | Point-prevalence (conducted over 2 weeks) | Residents at 69 nursing homes in Barcelona (Spain) who had not previously been diagnosed with COVID-19. | PCR |
| ONS 2020 (England, UK) | 10.7% | N/A | 293301 | 16 weeks | All care home residents in 9,081 surveyed homes in England (UK). No systematic testing. | Confirmed (no further details) |
| Brown 2020 (Ontario, Canada) | 6.6% | 5218 | 78607 | 7.5 weeks | All residents in Ontario (Canada) nursing homes with complete information (99% of all homes). No systematic testing. | Confirmed (no further details) |
| Rudolph 2020 (US) | 6.0% | 443 | 7325 | Point-prevalence | Veterans residing in VA community living centres (US) who participated in universal screening (excluding those who were tested due to symptoms prior to universal screening; those who were tested prior to admission to the centres; and those who were not tested). | PCR |
| Verbeek 2020 (The Netherlands) | 1.4% | 29 | 2011 | Unclear | Residents at 26 Dutch nursing homes included in the study (nationally representative for regions; no systematic testing). | Infected with COVID-19 (not further specified) |
| Mills 2020 c (Ohio, US) | 0.4% | 7 | 1794 | 14 weeks | People in assisted living facilities in Ohio (US) and receiving visiting medical care from the organisation behind the study (no systematic testing). | PCR |
| **Incidence of confirmed COVID-19 among long-term care staff (population-wide studies)** |  |  |  |  |  |  |
| Kennelly 2020 (Dublin and Eastern Ireland, Ireland) | 23.8% | 331 | 1392 | 12 weeks | Staff at nursing homes who responded to survey and provided staffing numbers (half of all responding nursing homes). | PCR |
| Borras-Bermejo 2020 (Barcelona, Spain) | 15.2% | N/A | 2,655 | Point-prevalence (conducted over 2 weeks) | Staff at 69 nursing homes in Barcelona (Spain) who had not previously been diagnosed with COVID-19. | PCR |
| Martin 2020 (Leon, Spain) | 9.5% | 7 | 74 | Point-prevalence study | Health care workers (GPs and nurses) working in 30 nursing homes in Leon (Spain) and who agreed to participate in the study. | Rapid diagnostic antibody test |
| ONS 2020 (England, UK) | 4.0% | N/A | 441,498 | 16 weeks | All care home staff in 9,081 surveyed homes in England (UK), including cleaning, catering and admin (no systematic testing). | Confirmed (no further details) |

Abbreviations: PCR, polymerase chain reaction

The incidence rate for cohorts of residents at long-term care institutions where an outbreak occurred varied widely. The lowest estimates was 0% over a three-week period and was observed in a South Korean long-term care hospital, where an infected care worker had been working throughout the facility for two days while symptomatic.^34^ Following the diagnosis of the index case, exposed care workers were quarantined at home, while remaining staff who continued to work were quarantined in a hotel. Considerably higher incidence rates of between 40.3% and 71.7% were reported from outbreaks in facilities in the US, UK, and France.

The incidence rate for cohorts of long-term care staff at outbreak facilities was overall lower compared to residents. Among nine studies testing all or close to all staff members, the rate of infections was generally below 10%, with the exception of one French study reporting a rate of 23.5% over six weeks (during weekly testing, no new people with COVID-19 were detected after the first two weeks) and a US study reporting a rate of 19.9% over six weeks across four outbreak facilities.^51,64^ Another point-prevalence study from the UK found that 4.1% of a sample of asymptomatic staff representing various roles across three nursing homes (including care workers as well as kitchen staff, administrators, and maintenance personnel) tested positive.^30^ The rate of infection was higher for the remaining three studies, but this included two where testing was only conducted for some staff members, and one report of an outbreak that did not provide details on testing.

Population-wide estimates of infection ranged from 0.4% to 40.8% for different populations of LTC users (Table 3). Only three studies were based on systematic testing, including the two studies reporting the highest rates of infection. However, some caveats about their findings should be noted. Kennelly et al. only included nursing homes in Dublin and Eastern Ireland that reported outbreaks and responded to their survey, thereby excluding more than half of all nursing homes in their sampling frame.^56^ Borras-Bermejo et al. tested residents and staff of 69 nursing homes in Barcelona but excluded those who already had a confirmed COVID-19 diagnosis.^52^ Rudolph et al. tested residents of Veteran Affairs community living centres in the US (predominantly male) but excluded those who had already been tested (due to showing symptoms, or prior to admission to a facility) and those who did not participate in universal testing.^66^

Two of the studies based on systematic testing also reported population-wide prevalence among LTC staff, with 23.8% of staff at nursing homes in Dublin and Eastern Ireland testing positive and 15.2% of staff at nursing homes in Barcelona.^52,56^ Another Spanish study found that 9.5% of health care workers in nursing homes in the region of Leon had antibodies, but was likely to underestimate true prevalence due to the testing strategy.^58^ Finally, a survey among facilities providing care to people living with dementia and older people in England found that 4.0% of staff members had contracted confirmed COVID-19 at some point during the pandemic (no systematic testing was in place at the time of the survey).^62^

Data on asymptomatic residents and staff who tested positive for COVID-19 was extracted from 13 studies (Table 4). ^22,26,30,31,35,36,39,40,48,51,52,56,63^ Definitions of symptomatic cases differed (see lists of symptoms in the table), as did the number of identified cases, leading to a range of asymptomatic people who contracted COVID-19 at the time of testing between 7% and 78% among long-term care residents, and between 24% and 100% among staff.

**Table 4:**
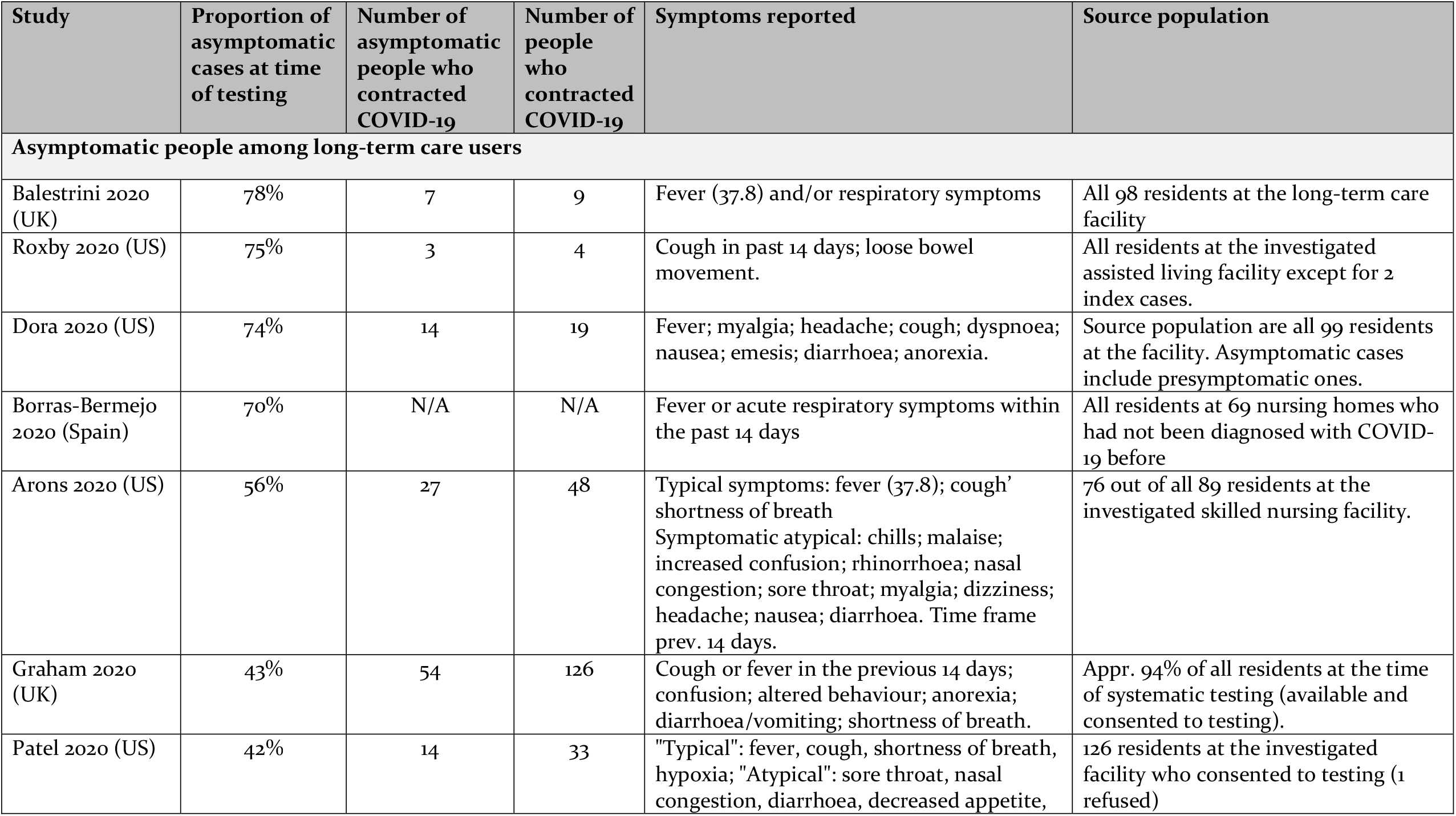
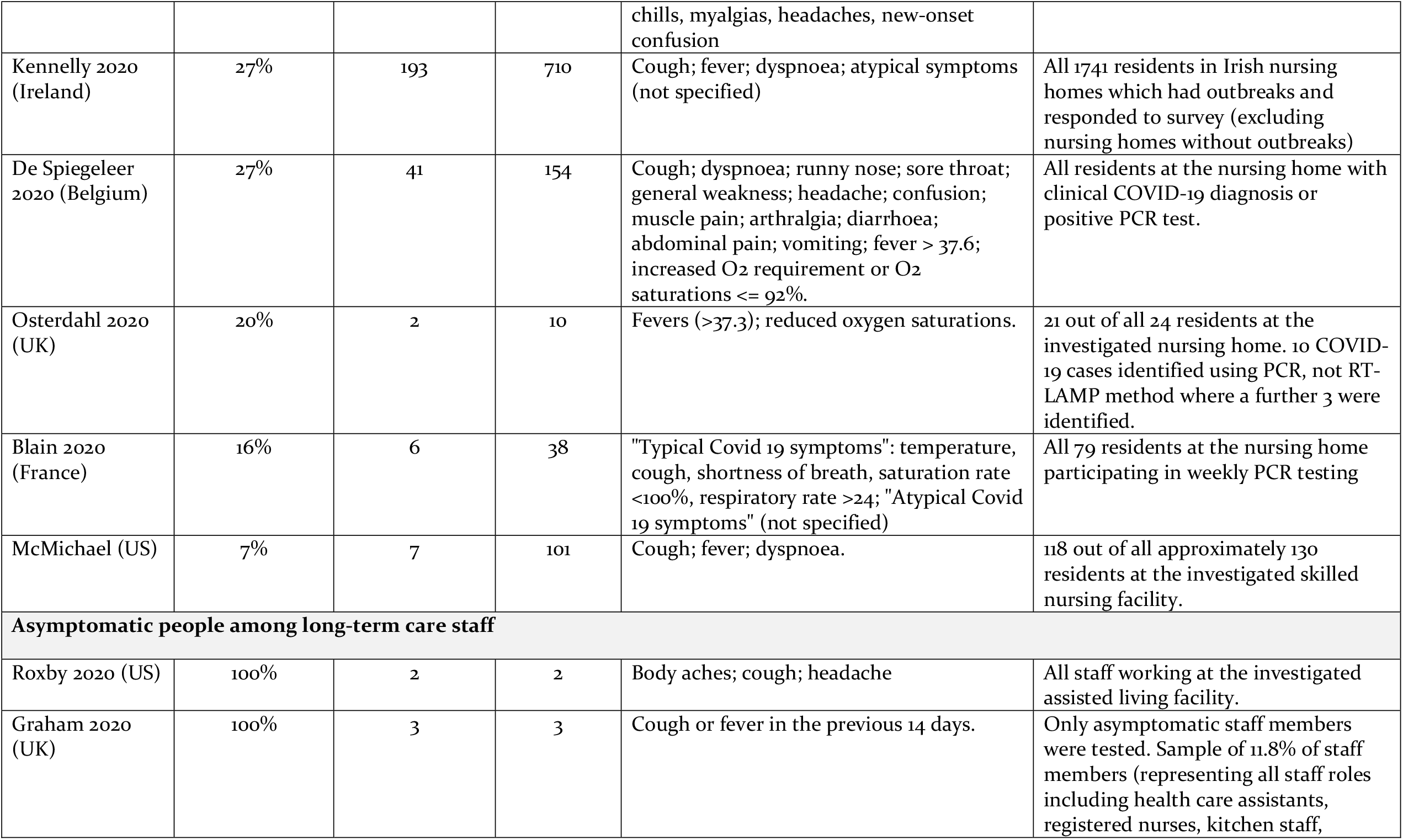
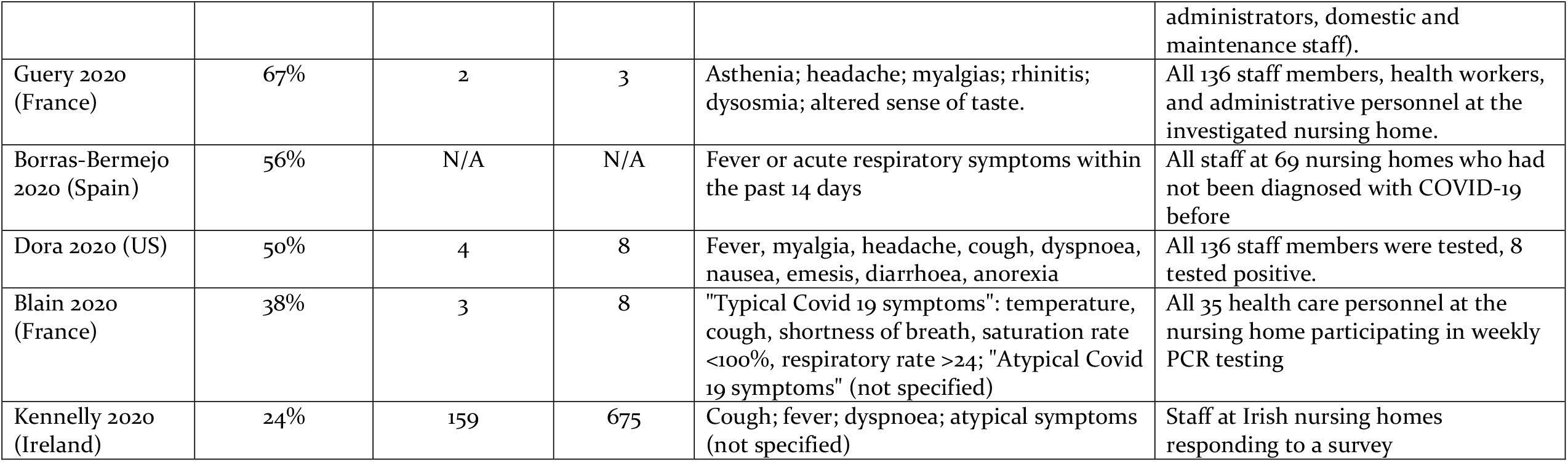
Proportion of asymptomatic cases at time of testing.

| Study | Proportion of asymptomatic cases at time of testing | Number of asymptomatic people who contracted COVID-19 | Number of people who contracted COVID-19 | Symptoms reported | Source population |
| --- | --- | --- | --- | --- | --- |
| **Asymptomatic people among long-term care users** |  |  |  |  |  |
| Balestrini 2020 (UK) | 78% | 7 | 9 | Fever (37.8) and/or respiratory symptoms | All 98 residents at the long-term care facility |
| Roxby 2020 (US) | 75% | 3 | 4 | Cough in past 14 days; loose bowel movement. | All residents at the investigated assisted living facility except for 2 index cases. |
| Dora 2020 (US) | 74% | 14 | 19 | Fever; myalgia; headache; cough; dyspnoea; nausea; emesis; diarrhoea; anorexia. | Source population are all 99 residents at the facility. Asymptomatic cases include presymptomatic ones. |
| Borras-Bermejo 2020 (Spain) | 70% | N/A | N/A | Fever or acute respiratory symptoms within the past 14 days | All residents at 69 nursing homes who had not been diagnosed with COVID-19 before |
| Arons 2020 (US) | 56% | 27 | 48 | Typical symptoms: fever (37.8); cough' shortness of breath<br>Symptomatic atypical: chills; malaise; increased confusion; rhinorrhoea; nasal congestion; sore throat; myalgia; dizziness; headache; nausea; diarrhoea. Time frame prev. 14 days. | 76 out of all 89 residents at the investigated skilled nursing facility. |
| Graham 2020 (UK) | 43% | 54 | 126 | Cough or fever in the previous 14 days; confusion; altered behaviour; anorexia; diarrhoea/vomiting; shortness of breath. | Appr. 94% of all residents at the time of systematic testing (available and consented to testing). |
| Patel 2020 (US) | 42% | 14 | 33 | "Typical": fever, cough, shortness of breath, hypoxia; "Atypical": sore throat, nasal congestion, diarrhoea, decreased appetite, chills, myalgias, headaches, new-onset confusion | 126 residents at the investigated facility who consented to testing (1 refused) |
| Kennelly 2020 (Ireland) | 27% | 193 | 710 | Cough; fever; dyspnoea; atypical symptoms (not specified) | All 1741 residents in Irish nursing homes which had outbreaks and responded to survey (excluding nursing homes without outbreaks) |
| De Spiegeleer 2020 (Belgium) | 27% | 41 | 154 | Cough; dyspnoea; runny nose; sore throat; general weakness; headache; confusion; muscle pain; arthralgia; diarrhoea; abdominal pain; vomiting; fever > 37.6; increased O<sub>2</sub> requirement or O<sub>2</sub> saturations ≤ 92%. | All residents at the nursing home with clinical COVID-19 diagnosis or positive PCR test. |
| Osterdahl 2020 (UK) | 20% | 2 | 10 | Fevers (>37.3); reduced oxygen saturations. | 21 out of all 24 residents at the investigated nursing home. 10 COVID-19 cases identified using PCR, not RT-LAMP method where a further 3 were identified. |
| Blain 2020 (France) | 16% | 6 | 38 | "Typical Covid 19 symptoms": temperature, cough, shortness of breath, saturation rate <100%, respiratory rate >24; "Atypical Covid 19 symptoms" (not specified) | All 79 residents at the nursing home participating in weekly PCR testing |
| McMichael (US) | 7% | 7 | 101 | Cough; fever; dyspnoea. | 118 out of all approximately 130 residents at the investigated skilled nursing facility. |
| **Asymptomatic people among long-term care staff** |  |  |  |  |  |
| Roxby 2020 (US) | 100% | 2 | 2 | Body aches; cough; headache | All staff working at the investigated assisted living facility. |
| Graham 2020 (UK) | 100% | 3 | 3 | Cough or fever in the previous 14 days. | Only asymptomatic staff members were tested. Sample of 11.8% of staff members (representing all staff roles including health care assistants, registered nurses, kitchen staff, administrators, domestic and maintenance staff). |
| Guery 2020 (France) | 67% | 2 | 3 | Asthenia; headache; myalgias; rhinitis; dysosmia; altered sense of taste. | All 136 staff members, health workers, and administrative personnel at the investigated nursing home. |
| Borras-Bermejo 2020 (Spain) | 56% | N/A | N/A | Fever or acute respiratory symptoms within the past 14 days | All staff at 69 nursing homes who had not been diagnosed with COVID-19 before |
| Dora 2020 (US) | 50% | 4 | 8 | Fever, myalgia, headache, cough, dyspnoea, nausea, emesis, diarrhoea, anorexia | All 136 staff members were tested, 8 tested positive. |
| Blain 2020 (France) | 38% | 3 | 8 | "Typical Covid 19 symptoms": temperature, cough, shortness of breath, saturation rate <100%, respiratory rate >24; "Atypical Covid 19 symptoms" (not specified) | All 35 health care personnel at the nursing home participating in weekly PCR testing |
| Kennelly 2020 (Ireland) | 24% | 159 | 675 | Cough; fever; dyspnoea; atypical symptoms (not specified) | Staff at Irish nursing homes responding to a survey |

### Evidence on case fatality rates in institutionalised long-term care settings

CFR is the proportion of people with confirmed COVID-19 who die. CFRs for 11 studies of all people at long-term care institutions facing outbreaks (including COVID-19 positive and negative residents and staff) are summarised in Table 5,^10,22,26,29,30,36,39,41,48,51,63^ while CFRs for seven studies of COVID-19 positive populations are described further down.^23,25,32,38,60,67,68^

**Table 5:**
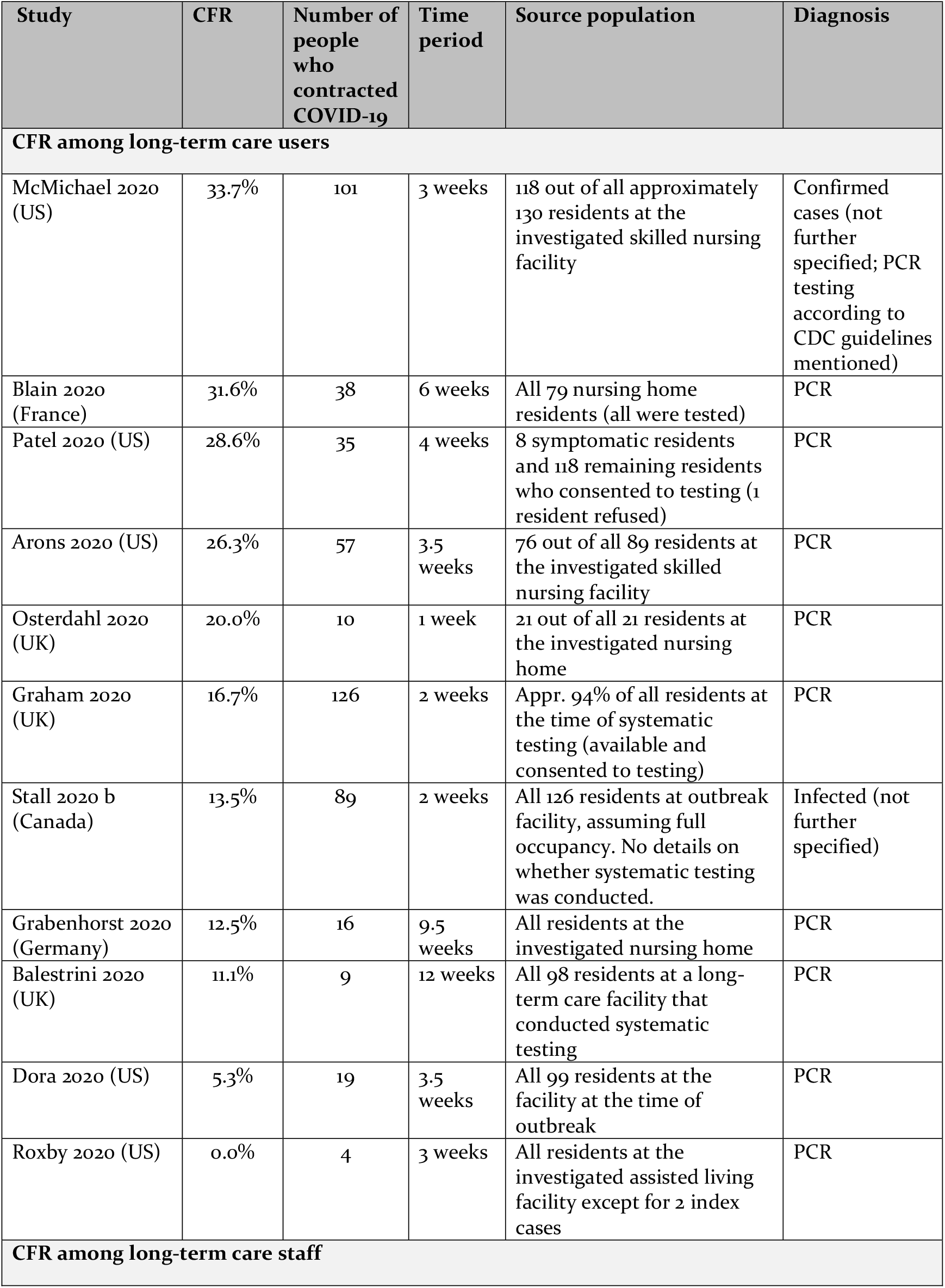
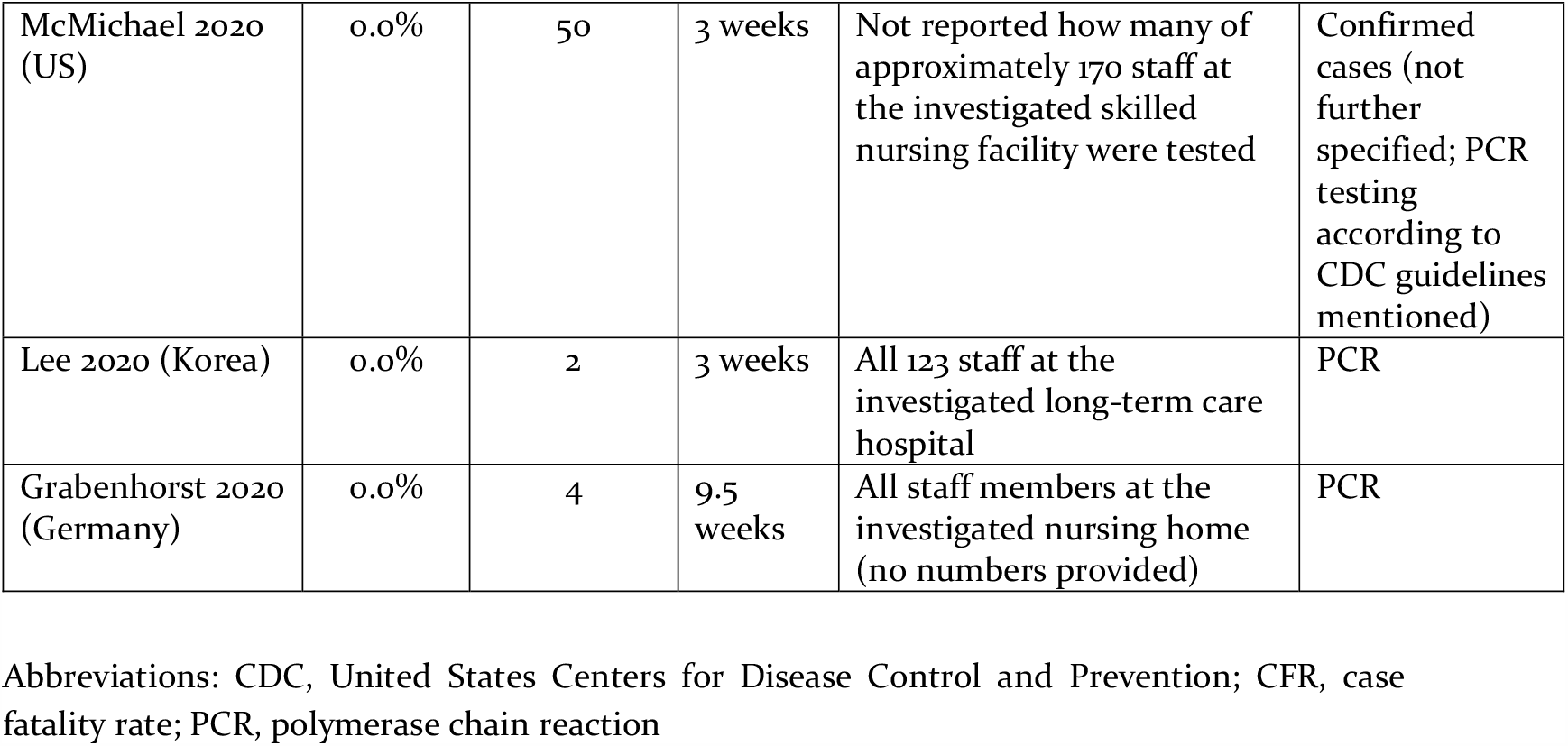
Case fatality rates among long-term care users and staff.

For most studies included in Table 5, the source population for the identification of people with COVID-19 were all or close to all (>90%) residents or staff at long-term care institutions where an outbreak occurred. The CFR among long-term care users for these studies ranged from 0% to 33.7%, with differences in follow-up time between one and 12 weeks. The CFR among long-term care staff was 0% in all included studies. These figures of outbreaks at individual facilities are complemented by a survey of nursing homes experiencing outbreaks in Dublin and Eastern Ireland, which found a CFR of 25.8% for residents with confirmed COVID-19.^56^

Not included in Table 5 are CFRs reported by seven studies for which the source population consisted exclusively of people who contracted COVID-19.^23,25,32,38,60,67,68^ Due to the absence of systematic testing, these studies tended to only include symptomatic people in the denominator, resulting in higher CFRs compared to studies based on systematic testing, as described below.

Prieto-Alhambra et al. found the 30-day mortality rate among 10,795 nursing home residents who were registered with a clinical or lab-confirmed COVID-19 diagnosis in a regional primary care database in Catalonia (Spain) to be 25.3% (95% CI 24.2-26.4%).^38^ This was considerably higher compared to the 30-day mortality rate for all other people with COVID-19 in the database (4.0%, 95% CI 3.9-4.2%), although these findings were not adjusted for age and underlying chronic conditions.

Baker et al. found that 40.7% of 60 nursing or residential home residents with lab-confirmed COVID-19 who were admitted to a teaching hospital in Newcastle (UK) died within a 28-day period.^23^ Compared to non-care home residents, the unadjusted odds ratio for death was 6.19 (95% CI 3.32-11.8).

De Smet et al. report that 52% of COVID-19 patients at a geriatric department were admitted from long-term care facilities, and the CFR for those was 28.6%.^25^

The source population was unclear for Kemenesi et al., which reported the number of deaths among all nursing home residents with confirmed COVID-19 in Hungary to be 4%.^32^ It is unclear how people contracting COVID-19 in nursing homes were identified.

Similarly, the CFR of 27.8% in Brown et al. was calculated across all nursing homes in the province of Ontario, Canada, but this was based on reporting of cases to the authorities, rather than on systematic testing of all residents.^68^

The CFR was 14.3% for seven people with confirmed COVID-19 among those receiving visiting medical care at assisted living facilities in Ohio (US).^60^

Finally, Verbeek et al. report that 16 of 29 people with confirmed COVID-19 at 26 nationally representative nursing homes in the Netherlands died, but did not provide a time period for these figures (CFR 55.2%).^67^

### Evidence on mortality rates in long-term care settings

Table 6 presents mortality rates for people who contracted COVID-19 from 11 studies of outbreaks at long-term care institutions.^26,29,30,34,36,39,41,48,51,54,63^ The source populations for these studies are all or close to all (>90%) residents or staff at long-term care institutions where an outbreak occurred. For these studies, the mortality rate for all or nearly all residents over a 1-to-12-week follow-up period was between 0.0% and 17.1%. Due to limited data on source population and causes of deaths, we did not include an outbreak report from another skilled nursing facility in the US in this Table.^50^ Assuming full occupancy at the 150-bed facility, and all 29 reported deaths having been caused by COVID-19, the mortality rate would be 19.3% of all residents over a 3.5-week period.

**Table 6:** COVID-19 mortality rates among long-term care users and staff.

| Study | Mortality rate of all users / staff | Number of users / staff | Number of people who contracted COVID-19 | Number of deaths among those who contracted COVID-19 | Time period | Source population |
| --- | --- | --- | --- | --- | --- | --- |
| **Mortality rate among long-term care users** |  |  |  |  |  |  |
| Diamantis 2020 (France) | 17.1% | 140 | N/A | 24 | 1 week | All residents at the long-term care facility. Note that number of deaths was "more than 24". No case definition was provided. |
| Blain 2020 (France) | 15.2% | 79 | 38 | 12 | 6 weeks | All 79 nursing home residents (all were tested) |
| Osterdahl 2020 (UK) | 9.5% | 21 | 10 | 2* | 1 week | 21 out of all 24 residents at the investigated nursing home |
| Stall 2020 b (Canada) | 9.5% | 126 | 89 | 12 | 2 weeks | All nursing home residents, assuming full occupancy of 126-bed facility |
| Patel 2020 (US) | 7.9% | 126 | 35 | 10 | 4 weeks | 8 symptomatic residents and 118 remaining residents who consented to testing (1 resident refused) |
| Graham 2020 (UK) | 6.7% | 313 | 126 | 21 | 2 weeks | Appr. 94% of all residents at the time of systematic testing (available and consented to testing) |
| Grabenhorst 2020 (Germany) | 1.6% | 122 | 16 | 2 | 9.5 weeks | All 122 residents at the nursing home at the time of systematic testing |
| Dora 2020 (US) | 1.0% | 99 | 19 | 1 | 3.5 weeks | All 99 residents at the facility at the time of outbreak |
| Balestrini 2020 (UK) | 1.0% | 98 | 9 | 1 | 12 weeks | All 98 residents at a long-term care facility that conducted systematic testing |
| Lee 2020 (Korea) | 0.0% | 193 | 0 | 0 | 3 weeks | All 193 inpatients at the investigated long-term care hospital who were exposed to an infected care worker |
| Roxby 2020 (US) | 0.0% | 80 | 4 | 0 | 3 weeks | All residents at the investigated assisted living facility except for 2 index cases |

| Mortality rate among care home staff |  |  |  |  |  |  |
| --- | --- | --- | --- | --- | --- | --- |
| Lee 2020 (Korea) | 0.0% | 123 | 2 | 0 | 3 weeks | All 123 staff at the investigated long-term care hospital |
| Grabenhorst 2020 (Germany) | 0.0% | 122 | 4 | 0 | 9.5 weeks | All 122 staff at the nursing home at the time of systematic testing |

\* Deaths caused by COVID-19 only. There was one further death among people who contracted COVID-19, but this was ascribed to a different cause.

**Table 7:** Incidence of hospitalisations among long-term care users and staff with COVID-19 diagnosis.

| Study | Incidence rate of hospitalisations | Number of people who contracted COVID-19 | Time period | Source population |
| --- | --- | --- | --- | --- |
| **Hospitalisations among long-term care users** |  |  |  |  |
| McMichael 2020 (US) | 54.5% | 101 | 3 weeks | All residents at the investigated skilled nursing facility who were confirmed cases (not further specified; PCR testing according to CDC guidelines mentioned) |
| Mills 2020 c (US) | 42.9% | 7 | 14 weeks | Confirmed cases among people receiving visiting medical care (no systematic testing) |
| Patel 2020 (US) | 37.0% | 35 | 4 weeks | Confirmed cases among 126 residents tested at outbreak facility |
| Yanover 2020 (Israel) | 34.4% | 67 | Not reported | Patients covered by Israeli health plan who had a SARS-CoV-2 positive PCR test and who were nursing home residents |
| De Spiegeleer 2020 (Belgium) | 24.0% | 154 | 6.5 weeks | All residents at the nursing home with clinical COVID-19 diagnosis or positive PCR test |
| Arons 2020 (US) | 19.3% | 48 | 3.5 weeks | All residents at the investigated skilled nursing facility with a positive PCR test |
| Prieto-Alhambra 2020 (Spain) | 16.1% | 10795 | 4 weeks | All patients included in a Catalan primary care database who are nursing home residents and have a clinical COVID-19 diagnosis or positive PCR test |
| Grabenhorst 2020 (Germany) | 12.5% | 16 | 9.5 weeks | All residents with a positive PCR test, identified through systematic testing of all residents |
| Roxby 2020 (US) | 0.0% | 4 | 3 weeks | All residents at the investigated assisted living facility with a positive PCR test except for 2 index cases |
| **Hospitalisations among long-term care staff** |  |  |  |  |
| McMichael 2020 (US) | 6.0% | 50 | 3 weeks | All health care personnel at the investigated skilled nursing facility who were confirmed cases (not further specified; PCR testing according to CDC guidelines mentioned) |
| Arons 2020 (US) | 0.0% | 26 | 3.5 weeks | All full-time staff members at the investigated skilled nursing facility with a positive PCR test |

There were two studies reporting on mortality among all staff members who were screened, and in both of these outbreaks, no member of staff had died after follow-up periods of 3 and 9.5 weeks, respectively.^29,34^

Information on excess deaths among long-term care residents was only available from two studies in the London area (UK). One study of outbreaks in four nursing homes estimated an increase in all-cause mortality by 203% for a two-month period compared to the average of the preceding two years.^30^ In contrast, a study of three different homes in the London area found the number of deaths over 12 weeks comparable to average mortality rates from the previous five years.^48^

Three studies reported the number of COVID-19 related deaths among wider populations of long-term care users. The proportion of all nursing home residents who died having contracted COVID-19 was 1.8% across all nursing homes in Ontario, Canada (1,452 deaths among 78,607 residents),^68^ and 0.8% across 26 nursing homes representing all regions in the Netherlands (16 deaths among 2,011 residents).^67^ However, neither of the two studies was based on systematic testing of residents and both could have underestimated the true number of residents dying having contracted the disease. A considerably higher proportion was reported for a sample of 21 nursing homes in Eastern Ireland and Dublin (10.5% of all residents over a 12-week period).^56^ However, this study was limited to nursing homes with active outbreaks and was missing information from approximately one third of homes in the sampling frame.

### Evidence on hospitalisations and ICU admissions from institutionalised long-term care settings

Nine studies provided information on the rate of hospitalisations among long-term care residents with COVID-19 diagnosis,^10,22,29,38,39,40,43,60,63^ (Table 6). Hospitalisation rates for long-term care residents varied between 0.0% and 54.4% for follow-up periods of between three and 14 weeks.

Hospitalisation rates for long-term care staff with COVID-19 diagnosis were 0.0% and 6.0% in two studies in US skilled nursing facilities.

Two studies reported the number of people who contracted confirmed COVID-19 among long-term care users who were admitted to the ICU. Arons et al. report that 5.3% of 48 nursing home residents with a positive PCR test were admitted to an ICU over a 3.5-week period.^22^ Roxby et al. report that none of the four residents at an assisted living facility with positive PCR test were admitted to an ICU over a three-week period.^39^

### Evidence on impact of COVID-19 on people who use long-term care community services

Only three included studies focused on people receiving long-term care in the community.

One US study reported on people with intellectual and developmental disability receiving long-term care services in the community, including in their family homes, foster care homes, or group homes (although some also lived in intermediate care facilities).^46^ Among a total population of 11,540 individuals, there were 66 with confirmed COVID-19 (0.6%) over a 100-day period. Only symptomatic people were tested. The CFR among people with confirmed COVID-19 was 4.5%, and 22.7% required hospitalisation.

The same organisation providing services to people with intellectual and developmental disability also reported on their experience providing home health and personal care to older people.^61^ Over 100 days, 67 people who contracted confirmed COVID-19 were detected (less than 0.3% of all clients). 47 of 67 were detected while living in the community. Among these, 17 required hospitalisation and 13 died.

The third study reported a total of 84 people with confirmed or suspected COVID-19 among the users of a memory unit and day care centre for people with cognitive disorders in Barcelona (Spain), with a CFR of 44.1%.^49^

### Evidence on outcomes in long-term care residents compared to others

Seven of the included studies compared outcomes in people who contracted COVID-19 between long-term care users and others. These studies generally found that long-term care users had worse outcomes, including higher 28-day-mortality (unadjusted odds ratio for death of nursing home or residential home residents admitted to hospital compared to non-residents: 6.19, 95% CI 3.32-11.8),^23^ 30-day-mortality (25.3%, 95% CI 24.2-26.4%, among nursing home residents who contracted COVID-19 and were registered in a primary care database compared to 4.0%, 95% CI 3.9-4.2%, among all other people with COVID-19 in the database),^38^ and overall mortality (incidence rate ratio for COVID-19 mortality comparing Ontario long-term care residents to community-living adults 70 years and older: 13.1, 95% CI 9.9-17.3),^27^ as well as increased risk of complicated disease (odds ratio for deteriorating disease, admission to ICU, or death, comparing nursing home residents to non-residents over 65 years of age: 2.48, 95% CI 1.29-4.65).^43^ Bhatraju et al. report that, among 24 patients admitted to the intensive care units of nine hospitals in the Seattle area (US), six (25%) were residents of skilled nursing facilities.^7^

Two studies did not find a statistically significant association between long-term care users and worse COVID-19 outcomes. De Smet et al. found that short-term mortality was not associated with long-term care residence in a cohort of COVID-19 patients at a geriatric department.^25^ Palaiodimos et al. did not find that nursing home residents fared worse than community-dwelling patients in a retrospective cohort study of the first 200 lab-confirmed COVID-19 cases in a teaching hospital in New York, US.^37^ There was no statistically significant difference between community-based and skilled nursing facility based patients for in-hospital mortality (OR 0.90, 95% CI 0.42-1.91; p= 0.779), increasing oxygen requirements (OR 1.14, 95% CI 0.59-2.21; p= 0.701), and intubation (1.39, 95% CI 0.59-3.27; p= 0.446).

### Evidence on burden of disease in the long-term care sector

Complementing studies of individual outbreaks, 14 studies provided evidence on the extent to which long-term care users are affected by the COVID-19 pandemic. These studies varied widely in their sampling frame (ranging from nationwide figures to single-centre case series of COVID-19 patients), and findings therefore need to be viewed in this context.

Kemenesi et al. report that in Hungary, as of 18 April 2020, 11% of all people with laboratory-confirmed COVID-19 in the country came from social homes, all of which were adult nursing homes.^32^ Similarly, Raciborski et al. report that 13.3% of all people with laboratory-confirmed COVID-19 in Poland up to 30 April 2020 were in nursing homes.^65^ Prieto-Alhambra et al. found that 8.9% of 121,263 people with COVID-19 registered in primary care records in Catalonia (Spain) were nursing home residents.^38^

Brown et al. found that 6.6% of all nursing homes in Ontario (Canada) had at least one person who contracted COVID-19 between 29 March and 20 May 2020.^42,68^ 86% of cases were concentrated in only 10% of nursing homes.

Li et al. also found infections in nursing homes in Connecticut (US) to be concentrated.^57^ 50% of 215 surveyed nursing homes reported any person who contracted COVID-19. While the average number of infected people was eight per home, 29% of homes reported more than ten. Lower staffing levels, higher quality ratings, and higher concentrations of Medicare or ethnic minority residents were predictive of higher numbers of infected people. Similarly, He et al. found that 35% of nursing homes in California (US) had at least one person who contracted the disease, with nursing home ratings and proportion of residents from ethnic minority groups predictive of COVID-19 infections and deaths.^55^ Across the US, Abrams et al. found that 31.4% of 9,395 surveyed nursing homes had at least one documented person with COVID-19.^19^ Mills et al. report that 1.3% of all homes for people with intellectual and developmental disability in the US supported by their organisation (including community-based sites, such as family and foster care homes, as well as institutional homes) had at least one person with confirmed COVID-19 over a 100-day period, although testing was limited to symptomatic individuals.^46^

The Office for National Statistics (ONS) for England conducted a large survey and found that 56% of all care homes catering to people living with dementia and older people had at least one person who contracted confirmed COVID-19.^62^ Higher levels of infection among residents were associated with prevalence of infection among staff, the use of bank or agency nurses, and different regions. In line with this finding about regional variation, Brainard et al. report that only 25 of 248 care homes in Norfolk (UK) had any people who contracted COVID-19.^53^ Detection of any cases was associated with the number of staff not directly involved in personal care.

Cabrera et al. report the results of systematic testing of all residents and staff in care homes in Galicia (Spain) and found the prevalence of confirmed COVID-19 to be 3.4% (no breakdown of these figures by long-term care users and staff was provided).^44^ 263 of 306 care homes did not have a single person with confirmed COVID-19.

Kim & Jiang found that three of the 12 largest clusters in South Korea were related to long-term care facilities, including two nursing homes and one psychiatric ward of a long-term care hospital.^33^ Das and Gopalan found that 46 out of 3,299 (1.4%) patients with confirmed COVID-19 in South Korea from 20 January to 30 April 2020 had been exposed at nursing homes (no information about whether these were residents, staff, or visitors).^24^

Gold reports that 20 of 305 (6.6%) of all hospitalised patients with laboratory confirmed COVID in Atlanta and Southern Georgia (US) were residents in a long-term care facility (study period: 1 to 30 March 2020).^28^ Also reporting on a cohort of hospitalised patients, Martin-Jimenez et al. found that 16.3% of deceased COVID-19 patients at their hospital in Madrid (Spain) were nursing home residents.

## Discussion

We report updated findings of a living systematic review of the spread of COVID-19 and outcomes in long-term care settings. Our findings based on review of 49 studies can be summarised as follows. First, outbreak reports and studies of wider populations of long-term care users showed the severe impact of the pandemic on this group. Outbreaks at long-term care facilities can affect more than thirds of residents and lead to the deaths of a little under one fifth of residents. Excess risk of severe outcomes for long-term care users after contracting COVID-19 was also found in several studies (including increased risk of death), although not all studies accounted for case mix, and other studies did not find increased risk for long-term care users. Second, included studies showed substantial variation in how widely the disease spread among both residents and staff, and how many residents died as a result of COVID-19 outbreaks in long-term care facilities. While it is currently unclear what is driving the variation in spread of disease and outcomes, some outbreaks have been contained successfully, suggesting that future research should explore the source of this variation to provide urgently needed evidence to better manage COVID-19 in long-term care. Some of the included studies also provided early evidence on characteristics of nursing homes predictive of higher numbers of people contracting COVID-19, which will need to be substantiated through future research. Evidence on impact of COVID-19 on long-term care in the community is still scarce, even though this represents a group that is potentially highly vulnerable to infection (as they rely on care from others) and at risk of severe outcomes.^69^ Third, a substantial proportion of people with COVID-19 detected during systematic screening of residents (as many as 75%) and staff (up to 100%, although case numbers were very low) of long-term care facilities were asymptomatic at the time of testing, casting doubts over the appropriateness of symptoms-based strategies in this setting. Finally, reporting standards of included studies were variable and often poor, highlighting the need to harmonise research practices and reporting standards in this body of fast-evolving literature.

### Impact of COVID-19 on the long-term care sector

The findings of this living systematic review underline the urgent need for decisive policy action to tackle the COVID-19 pandemic in the long-term care sector. The combination of older, chronically multimorbid people, living in close proximity to each other has contributed to this population being particularly vulnerable to the COVID-19 pandemic. This vulnerability has been mirrored in official figures which show that deaths in long-term care users now make up more than 50% of all COVID-19 related deaths in at least five countries, and more than 30% in 16 of 19 countries reporting relevant data.^8^

Emerging evidence summarised in this review also shows potentially excessive risk of severe outcomes, including a higher risk of death, among long-term care residents compared to non-long-term care residents of similar age. While official deaths data for care homes in most countries only includes people who either tested positive or had COVID-19 mentioned in the death certificate, data from England and Wales shows that the number of excess deaths of care home residents during the pandemic (compared to the number of deaths in the same period in previous year) was almost double the number of deaths that had been registered as being linked to COVID-19.^8^**Error! Bookmark not defined**. This suggests that current official estimates of the mortality impact of COVID-19 in care homes in most countries may underestimate the full impact of the pandemic, be it because of lack of attribution of deaths to COVID, or because of other indirect effects such as reduced access to usual health care for non-COVID conditions.

This evidence highlights the need to develop targeted policies to both prevent outbreaks in long-term care settings, and to manage them effectively once they occur. In many countries, long-term care was not a priority in the early stages of the pandemic. In the UK, whilst policymakers had been aware of this risk early on in the pandemic,^70^ inadequacies in the testing strategy and a focus on ensuring bed capacity in the secondary care sector is likely to have undermined mitigation of the spread in care homes. Until 16 April 2020, three days after the peak in daily deaths, it was still possible for UK hospitals to send residents back to their care homes without having to test them for COVID-19.^71^

However, policymakers are increasingly aware of the scale of the problem in long-term care and starting to develop responses. For example, the WHO European Region Office has developed a list of ten policy objectives to tackle COVID-19 in long-term care, starting with the maintenance of long-term care services during the pandemic.^72^ This was recently expanded and updated by the WHO.^73^ Individual countries have developed their own set of policy responses, including implementing national task forces to coordinate responses in long-term care, the use of disease surveillance tools to monitor outbreaks in care homes and deployment of rapid response teams to manage them, reducing occupancy in care homes, and policies to increase the number of available staff.^74^ Other responses were aimed at preventing the disease entering care homes, including isolation of care home residents, restrictions or banning of visits, measures to reduce the risk of disease spreading through staff, and quarantining of residents discharged from hospital upon re-entering the care home. Importantly, as the pandemic continues over a prolonged period, attention will need to be paid to ensure continuing care and maintaining the health and wellbeing of both long-term care users and providers.

### Variation in infection rates and outcomes across countries and individual facilities

This review has shown considerable variation in the number of long-term care users and staff who contract the disease after an outbreak in a facility. In some cases, more than half of the resident population was infected. In other cases, outbreaks were contained to low numbers or even preventing a single confirmed infection among residents. Included studies were not designed to test the effectiveness of different strategies to prevent or contain outbreaks, leaving open questions about the factors driving the observed variation. Possible explanations for comparatively low infection rates in individual outbreaks include decisive action to isolate potentially infected staff members and removing people with confirmed COVID-19 from the facility,^34^ cohorting of infected residents,^26,29^ weekly serial facility-wide testing,^26^ as well as hygiene measures and comparatively spacious and more spread-out residents in an assisted living facility (compared to a nursing home).^39^ We aim to examine these factors in more detail in a living systematic review of COVID-19 interventions in long-term care parallel to this one. In addition to evaluating the effectiveness of different strategies in containing outbreaks, their impact on the wellbeing of long-term care users and staff should be assessed.

It will be important to design such studies with scientific rigour in order to provide meaningful and generalisable evidence to guide decision making. The case of experimental administration of post-exposure prophylaxis hydroxychloroquine for patients and staff at a long-term care facility in South Korea highlights the need for methodologically robust studies. In the South Korean example, lack of a control group made it impossible to attribute the success in containing the outbreak (no patient and only one staff member other than the index case were infected over a 2-week period) to post-exposure prophylaxis.^34^ In the meantime, a randomised controlled trial was published and showed no efficacy of hydroxychloroquine prophylaxis after exposure to COVID-19,^75^ making it appear more likely that strict isolation measures put in place at the South Korean facility contributed to containing the virus.

A strategy that has increasingly attracted attention is systematic screening of residents and staff at affected facilities. Our review underlines the importance of diagnostic testing as compared to symptoms-based screening. Several included studies reported the number of infected people detected through RT-PCR testing who were asymptomatic at the time. Due to the range in the number of people who contracted COVID-19 in these studies (4-710) it was difficult to infer a reliable proportion of those residents that were asymptomatic and yet were found to be COVID-19 positive on RT-PCR testing, but included studies suggest that this could be a substantial minority or even the majority of infected residents (range of asymptomatic cases among residents at time of testing, 7-78%). For care home staff, the small numbers and sampling methods to identify people with confirmed COVID-19 made it impossible to make robust inferences about the numbers of asymptomatic employees. Further information about this would warrant a more systematic testing strategy across all care home workers and residents. Indeed, such nationwide comprehensive testing of the care home population including staff is being conducted in Belgium, showing that 74% of residents who contracted COVID-19 and 76% of staff who contractred COVID-19 were asymptomatic at the time of testing.^76^

Some of the included studies also highlighted that people with asymptomatic infections may develop symptoms within a period of about one week.^10,22,26^ Future studies should plan to follow up identified people with COVID-19 to better understand symptoms and apply a more robust definition of what constitutes symptomatic. Whilst there is seemingly broad agreement with regards to respiratory symptoms, two studies did not consider gastrointestinal symptoms (anorexia, diarrhoea, abdominal pain, vomiting) within their definition of symptomatic. Consensus may also be needed on a definition of fever; it was interesting to note that two of the included studies in this review defined fever at 37.3 and 37.8, respectively.^22,36^

### Improvements to reporting of outbreaks

Lack of common standards in the reporting of outcomes has long been recognised as a major challenge for synthesising research findings.^77^ In this systematic review, substantial differences across the included set of studies precluded a quantitative synthesis of results. Studies differed in how testing was conducted (comprehensive testing vs. convenience samples; varying time periods over which outcomes data was collected, including infections). We were also unable to ascertain the homogeneity of different populations due to a lack of reporting of their characteristics. For example, some of the outbreak investigation reports failed to report characteristics of long-term care residents, such as mean age, sex distribution, comorbidities, and ethnicity. In other studies, it was sometimes unclear whether all long-term care users or staff in the sample frame had been tested, and how testing was conducted. These limitations highlight the need to establish minimum reporting standards for future studies evaluating COVID-19 related mortality and spread of disease in LTC settings.

### Limitations

This living review had some limitations. First, we extracted the number of people who contracted COVID-19 as defined by study authors when there was no specific confirmatory diagnostic test mentioned in the study, which may have overestimated the number of people with confirmed COVID-19. Second, we also relied on the definitions used by study authors for deaths due to COVID-19. These sometimes relied on official mortality figures, which share the limitations of the underlying data sources. Third, we report the proportion of long-term care users who were hospitalised due to COVID-19 but this is not necessarily an indicator for severity of disease, as it is likely to partially reflect differences in policies for transferring patients to acute care hospitals. Fourth, we deviated from our protocol due to the unanticipated large volume of research identified in this area. Instead of completing all review steps in double, one reviewer was responsible for study inclusion and data extraction. However, we implemented broad eligibility criteria in order to ensure no relevant studies were missed, and all studies deemed eligible for inclusion were reviewed by the same reviewer to ensure consistency.

### Conclusions

Long-term care users are particularly vulnerable during the COVID-19 pandemic, facing substantial risk of infection and death. Outbreak reports from individual long-term care facilities have shown wide variation in the spread of disease and outcomes among residents and staff. Further research into the factors determining successful prevention and containment of COVID-19 outbreaks in long-term care is needed, including for institutional and community-based services.

## Data Availability

Raw data are presented in the manuscript. Additional data available from authors upon request.

## Notes

### Competing Interest Statement

The authors have declared no competing interest.

### Clinical Protocols

https://www.crd.york.ac.uk/prospero/display_record.php?RecordID=183557

### Author Declarations

This is a systematic review using published summary-level data.

### Summary of Updates

This is the second update of a living systematic review (first published on 9 June 2020). Since the last update, 21 new studies were included.

